# Ischemic Stroke Complicated by Acute Decompensated Heart Failure and Its Association with Hemoglobin Levels

**DOI:** 10.1101/2024.12.30.24319796

**Authors:** Yusuke Yamazaki, Yasuyuki Shiraishi, Shun Kohsaka, Shogo Ikegami, Takashi Kohno, Yuji Nagatomo, Mitsunobu Kitamura, Munehisa Sakamoto, Michiru Nomoto, Atsushi Mizuno, Toshiyuki Takahashi, Masatake Kobayashi, Satoshi Higuchi, Masaki Ieda, Tsutomu Yoshikawa, the West Tokyo Heart Failure Registry Investigators

**Author notes:** Address for Correspondence: Yasuyuki Shiraishi, MD, PhD Department of Cardiology, Keio University School of Medicine, 35 Shinanomachi Shinjuku-ku, Tokyo, Japan, 160-8582.

## Abstract

**Background:** Ischemic stroke (IS) is a serious complication in heart failure, particularly following hospitalization for acute decompensated heart failure (ADHF). However, its actual incidence during the acute and stable treatment phases remains unclear. Moreover, the association between IS and hemoglobin levels, which fluctuate during these phases, has not been comprehensively investigated.

**Methods:** Patient-level data from 2018 to 2024 were extracted from a prospective multicenter cohort study that consecutively enrolled patients hospitalized with ADHF. Cerebrovascular events were confirmed by local board-certified neurologists using imaging modalities. The incidence of IS and its association with hemoglobin levels, categorized by the universal anemia definition (hemoglobin <13.0 g/dL for men and <12.0 g/dL for women) were examined in early (within 30 days) and late phases (beyond 30 days) following ADHF hospitalization. Fine–Gray models were used for early IS analyses, and Cox proportional hazards models with time-varying covariates (hemoglobin levels) were applied for late IS. Results: A total of 5,106 patients (median age 79 years; men 58%) were analyzed, and 115 (2.3%) developed IS over a median follow-up of 13 months. The incidence was higher in the early phase (median onset 7 days, *p for trend*=0.002). Cardioembolic stroke (61.7%) was the predominant subtype throughout phases. After covariate adjustments, higher hemoglobin levels (non-anemia) were associated with increased risk of early IS (sHR 2.05,95%CI 1.09– 3.83, p=0.03), but not with late IS (adjusted HR 1.21,95% CI 0.65–2.23, p=0.55), with restricted cubic spline further demonstrating non-linear phase-dependent differences in the impact of hemoglobin levels on IS risk.

**Conclusion:** IS occurred across phases of ADHF management, with higher incidence within 30 days after hospitalization. The relationship between hemoglobin levels and IS risk varies by phase, highlighting non-anemia as a potential marker for high-risk patients in the early phase. Incorporating hemoglobin levels into risk stratification could guide targeted screening and preventive strategies.

## Introduction

Heart failure (HF) is a leading cause of mortality worldwide, affecting over 64 million people.^1^ Ischemic stroke (IS) is a serious complication in HF, associated with increased mortality, substantial functional decline, reduced quality of life, and significant healthcare costs.^2–4^ HF is a recognized risk factor for IS,^5–7^ and previous studies have demonstrated that patients with reduced left ventricular ejection fraction (LVEF) have a substantial risk of IS, with annual incidences ranging from 1.5% to 2.7%.^8–10^ Interestingly, recent research has highlighted that IS risk remains comparable across a broad spectrum of LVEF levels.^11,12^ However, HF is not a singular, static condition; patients’ HF status can differ significantly, particularly after acute decompensated heart failure (ADHF).^13^ Given that numerous hazardous events can occur after the initial stabilization of ADHF, it is crucial to identify the vulnerable periods for IS in these patients.

Evidence for effective risk stratification and preventive interventions for IS in HF patients without atrial fibrillation (AF) remains limited, underscoring the need for future research^14–16^. One of the challenges in post-ADHF care is the substantial fluctuation in fluid status during the treatment period, which is partially reflected in hemoglobin levels.^17,18^ Assessing the association between hemoglobin levels and IS in ADHF patients may help identify phases during which these patients are particularly vulnerable to thromboembolic events.

In this study, we used data from a prospective multicenter ADHF cohort to assess the incidence of IS and its association with hemoglobin levels post-ADHF hospitalization. We also aimed to identify high-risk periods for IS following ADHF as this cohort involves long-term follow-up information, providing advantages for temporal analysis.

## Methods

The data supporting the findings of this study are available from the corresponding author upon reasonable request.

### Study Cohort

The design of the West Tokyo Heart Failure Registry-2 (WET-HF2) has been previously described.^19–21^ Briefly, the WET-HF2 is a prospective, multicenter cohort study designed to collect data on the clinical backgrounds and outcomes of consecutive patients with ADHF who were hospitalized for urgent treatment within eight tertiary care hospitals in the Tokyo area, Japan. When compared with other ADHF cohorts, WET-HF2 stands out as one of the most representative of ADHF population in Japan.^20,22^ Patients presenting with acute coronary syndrome or those aged <20 years were not included in this database. Individual cardiologists at each institution made the clinical diagnosis of ADHF based on the signs and symptoms of HF and the level of natriuretic peptides (i.e., B-type natriuretic peptide [BNP] ≥100 pg/mL or N-terminal pro-B-type [NT-pro BNP] ≥300 pg/mL) at the time of hospitalization.^23^ The patient data were collected by trained clinical research coordinators using a web-based electronic data capture system with a robust data query engine and system validations for data quality. The principal investigators (Y.S. and S.K.) performed periodic queries to verify the reporting data quality.

One-, two-, and five-year follow-up examinations were conducted on all patients via mail, phone interviews, and chart reviews after enrollment. Dedicated study coordinators updated the status of the major cardiovascular events (i.e., death, HF rehospitalization, major bleeding, stroke, etc.) and the procedures (i.e., device implantation, surgical or percutaneous operations, heart transplant, etc.).

Before launching the WET-HF2 registry, an objective and detailed design was provided for clinical trial registration with the University Hospital Medical Information Network (UMIN000032169). The study protocol was approved by the institutional review boards at each site, and the research was conducted in accordance with the Declaration of Helsinki. According to the Ethical Guidelines for Medical and Health Research Involving Human Subjects and Personal Information Protection Law in Japan, informed consent was obtained from each patient before the study began.

### Study Population

We enrolled 5,106 consecutive patients with ADHF in the WET-HF2 registry between April 2018 and April 2024. Patients were classified into two groups based on the presence of IS as a complication after ADHF hospitalization. Patients complicating with IS were further divided into the early-phase (within 30 days) and late-phase (beyond 30 days) groups, based on the timing of IS onset after ADHF hospitalization, as suggested by prior research that the risk of IS was highest within the first 30 days after discharge from HF hospitalization.^13^

### Variable Definitions

According to the World Health Organization criteria, anemia was defined as hemoglobin level <13 g/dL for men and <12 g/dL for women at hospitalization, according to the WHO scientific definition. When adding into multivariable models, high levels of natriuretic peptides (High NP) were defined as a BNP level >1000 pg/mL or an NT-pro BNP level >4000 pg/mL at hospitalization.^24^ The CHADS₂ score was calculated using proposed risk factors, including congestive heart failure, hypertension, age ≥75 years, diabetes mellitus, and previous stroke.^25^

Left ventricular ejection fraction (LVEF) was measured using the modified Simpson’s method, with reduced ejection fraction defined as LVEF ≤40%. Transthoracic echocardiography was performed during the index hospitalization in the compensated phase by highly experienced echocardiographers or cardiologists. The New York Heart Association (NYHA) functional classification was assessed at the time of hospitalization by the treating cardiologists at each institution. Estimated glomerular filtration rate (eGFR) was calculated using the Modification of Diet in Renal Disease Equation for Japanese Patients, as proposed by the Japanese Society of Nephrology.^26^

### Study Endpoint

The primary outcome of this study was the incidence of IS following an ADHF hospitalization. IS event was defined as the presence of new focal neurological symptoms lasting over 24 hours, supported by corresponding imaging findings including computed tomography and magnetic resonance imaging, and diagnosed by neurologists at each institution. The underlying cause of IS was classified according to the Trial of Org 10172 Acute Stroke Treatment (TOAST) classification ^27^ : cardioembolic, large-artery atherosclerosis, small-vessel disease, and stroke of undetermined cause.

### Statistical Analysis

We first divided patients into the IS and non-IS groups to compare their patient characteristics. We then conducted a comparative analysis of the patient characteristics between early-phase IS and late-phase IS groups. With respect to descriptive analyses, continuous variables are presented as median with interquartile range (IQR), while categorical variables are expressed as counts and percentages, using Mann-Whitney U tests for continuous variables and χ² tests or Fisher’s exact tests for categorical variables.

Next, we described the temporal trends in the incidence of IS during the entire period, early-phase, and late-phase, using the Jonckheere-Terpstra test. In addition, we analyzed the proportion of each etiology of IS based on the TOAST classification, by the entire period, early-phase, and late-phase.

To investigate the association between hemoglobin levels and time to IS, we performed phase-specific analyses. For overall- and early-phase IS, Fine and Gray models were employed to account the competing risk of death, yielding sub-distributional hazard ratios (sHRs) with 95% confidence intervals (CIs). For late-phase IS, Cox proportional hazards models with time-varying covariates were used to avoid immortal time bias and address violations of the proportional hazards assumption.^28^ Hemoglobin levels were treated as a time-dependent variable and measured at hospitalization, discharge, and one-year post-discharge. We tested the statistical significance of nonlinearity in the hemoglobin level by comparing the spline model with the linear model, ultimately finding a non-linear association between the hemoglobin level and IS. Therefore, we added the status of anemia as a binary variable into the models. The covariables included in the adjusted model were selected based on their potential to confound the association between hemoglobin levels and IS. To address the possibility of model over-fitting, given the relatively limited number of events compared to the number of covariates, we carefully selected covariates based on clinical relevance and prior literature. We ensured that the ratio of events to covariates met the recommended criteria to maintain the validity of the model estimates. These covariables in the overall model comprised established risk factors for IS, including age, sex, hypertension, diabetes mellitus, prior history of stroke, AF (defined as a previous history and electrocardiogram findings on presentation), high levels of natriuretic peptide, reduced LVEF (≤40%), and oral anticoagulants.^8,29,30^ In adjusted models for early-phase and late-phase IS, we selected CHADS2 score, sex, AF, high levels of natriuretic peptide, reduced LVEF, and oral anticoagulants, considering the number of IS incidence to avoid model-overfitting. Scaled Schoenfeld residuals were used to assess the proportionality assumption, which showed no violation. Furthermore, we employed the restricted cubic spline with 4 knots at the 5^th^, 35^th^, 65^th^, and 95^th^ centiles to evaluate the nonlinear relationship between hemoglobin levels and hazard ratio for IS, adjusting for potential covariates such as CHADS₂ score, sex, AF, high levels of natriuretic peptide, reduced LVEF, and oral anticoagulants.

To ensure the robustness of our findings, we conducted several sensitivity analyses. We stratified the patients based on LVEF (<40% vs. ≥40%) into two groups: reduced LVEF (<40%) and preserved LVEF (≥40%). Additionally, we stratified patients by the presence or absence of AF. For the time-to-event analysis of IS, we used Fine–Gray models to account for the competing risk of death from causes other than IS. In the LVEF-stratified analysis, sex, CHADS₂ score (which encompasses congestive heart failure, hypertension, age ≥75 years, diabetes mellitus, and prior stroke or transient ischemic attack), and oral anticoagulation were accounted as potential covariates. For the analysis stratified by AF presence, adjustments were performed for sex, reduced EF, and CHADS₂ score. Similarly, in the late-phase analysis, considering hemoglobin levels as a time-dependent covariate, we used time-varying Cox proportional hazards models. Furthermore, we explored the potential nonlinear relationship between hemoglobin levels and IS risk using restricted cubic spline functions within the Fine–Gray models. We selected knot positions based on percentiles of the hemoglobin distribution to capture possible nonlinear associations effectively. The spline models were adjusted for sex and CHADS₂ score.

Statistical analyses were performed using SPSS (version 29.0, SPSS Inc., Chicago, IL, USA) and R software (Version 4.4.1; R Foundation for Statistical Computing, Vienna, Austria) with the “cmprsk”, “rms”, and “survival” packages for competing risk and survival analyses, respectively. All tests were 2-sided, and P <0.05 was considered statistical significance.

## Results

### Patient characteristics

In the entire cohort, the median age was 79.0 years (IQR; 70.0–86.0), median LVEF was 45% (IQR; 31.0–58.0), and 58.2% of the patients were men. A history of prior HF hospitalizations was present in 27.1%, while 41.8% had AF. The median hemoglobin level was 12.0 (IQR: 10.4– 13.7) g/dL, with anemia present in 57.4%. The baseline characteristics based on presence or absence of IS are shown in **Table 1**. There were no significant differences between patients with and without IS regarding LVEF, AF, CHADS2 score, use of antiplatelets/anticoagulants, or hemoglobin levels. Notably, when compared to late-phase IS patients, those with early-phase IS tended to be younger, with non-ischemic cardiomyopathy as the underlying etiology including dilated and hypertrophic cardiomyopathies, higher heart rates at admission, less frequent use of loop diuretics and anticoagulants prior to hospitalization, and higher hemoglobin levels (**Table S1**).

**Table 1.** Patient characteristics with and without ischemic stroke

|  | Ischemic Stroke<br>(n = 115) | No Ischemic Stroke<br>(n = 4991) | p-value |
| --- | --- | --- | --- |
| Age, years | 79.0 [68.5–86.0] | 79.0 [70.0–86.0] | 0.76 |
| Men, n (%) | 69 (60) | 2092 (58.1) | 0.69 |
| Body mass index, kg/m<sup>2</sup> | 22.6 [20.4–25.3] | 23.0 [20.3–26.1] | 0.51 |
| Systolic blood pressure, mm Hg | 142 [119–162] | 138 [119–161] | 0.50 |
| Heart rate, per min | 96 [75–114] | 92 [74–112] | 0.49 |
| NYHA class III or IV, n (%) | 101 (87.8) | 4298 (86.1) | 0.70 |
| LVEF, % | 41.0 [30.5–58.0] | 45.0 [31.0–58.0] | 0.43 |
| LVEF ≤40%, n (%) | 54 (48.6) | 2055 (43.0) | 0.25 |
| Underlying etiology |  |  |  |
| Coronary artery disease, n (%) | 28 (24.3) | 1253 (25.1) | 0.94 |
| Valvular disease, n (%) | 30 (26.1) | 1183 (23.7) | 0.63 |
| Non-ischemic cardiomyopathy, n (%) | 22 (19.1) | 914 (18.3) | 0.92 |
| Others, n (%) | 30 (26.1) | 1390 (27.9) | 0.76 |
| Current smoker, n (%) | 12 (10.5) | 555 (11.1) | 0.94 |
| Prior HF hospitalization, n (%) | 22 (19.3) | 1354 (27.3) | 0.07 |
| Atrial fibrillation, n (%) | 42 (36.8) | 2080 (42.0) | 0.32 |
| Hypertension, n (%) | 74 (64.9) | 3300 (66.6) | 0.79 |
| Diabetes, n (%) | 41 (36.0) | 1556 (31.4) | 0.35 |
| Prior stroke, n (%) | 22 (19.3) | 687 (13.9) | 0.13 |
| COPD, n (%) | 5 (4.4) | 221 (4.5) | 1 |
| CHADS<sub>2</sub> score | 3 [2–4] | 3 [2–4] | 0.45 |
| BNP, pg/mL <sup>#</sup> | 681 [398–1575] | 709 [415–1258] | 0.89 |
| NT-proBNP, pg/mL <sup>#</sup> | 3784 [2499–10129] | 4555 [1190–7920] | 0.97 |
| eGFR, mL/min per 1.73m<sup>2</sup> | 48.4 [33.4–62.4] | 46.2 [31.5–60.8] | 0.45 |
| Hemoglobin, mg/dL | 12.2 [10.5–13.3] | 12.0 [10.3–13.7] | 0.83 |
| Anemia, n (%) <sup>\$</sup> | 67 (58.3) | 2863 (57.4) | 0.92 |
| Loop diuretics, n (%) | 49 (42.6) | 2271 (45.5) | 0.60 |
| Beta blocker, n (%) | 51 (44.3) | 2166 (43.4) | 0.91 |
| ACEi/ARB/ARNI, n (%) | 44 (38.3) | 1958 (39.2) | 0.33 |
| MRA, n (%) | 16 (13.9) | 891 (17.9) | 0.28 |
| SGLT2 inhibitor, n (%) | 8 (7.0) | 318 (6.4) | 0.95 |
| Antiplatelets, n (%) | 38 (33.0) | 1438 (28.8) | 0.38 |
| Anticoagulants, n (%) | 35 (30.4) | 1826 (36.6) | 0.21 |
| ICD, n (%) | 1 (0.9) | 139 (2.8) | 0.38 |
| CRT, n (%) | 0 (0) | 58 (1.2) | 0.64 |

<sup>#</sup> BNP was measured in 3393 patients and NT-pro BNP was measured in 1662 patients.
<sup>\$</sup> defined as hemoglobin level <13 g/dL for men and <12 g/dL for women.
Abbreviations: HF: heart failure, COPD: chronic obstructive pulmonary disease, NYHA: New York Heart Association, LVEF: left ventricular ejection fraction, ACEi: angiotensin-converting enzyme inhibitor, ARB: angiotensin receptor antagonist, ARNI: angiotensin receptor-neprilysin inhibitor, MRA: mineralocorticoid receptor antagonist, SGLT2: sodium-glucose cotransporter-2, ICD: implantable cardioverter defibrillator, CRT: cardiac resynchronization therapy

### Incidence, subtypes, and characteristics of ischemic stroke

Among 5,106 patients, 115 (2.3%) developed IS over a median follow-up of 13 months (IQR; 10-25) after hospitalization for ADHF (**Figure S1**), and an annual rate of IS was 3.6% per person. Patients with IS were categorized into early-phase (n = 56) and late-phase (n = 59) groups. In the early-phase group, IS events frequently occurred in the initial two weeks, with a median onset of 7 days (IQR; 3–12, *p for trend* = 0.02) (**Figure 1**). During the index hospitalization, IS occurred in 54 (1.1%) patients, where four cases were associated with new-onset AF after hospitalization.

**Figure 1.**
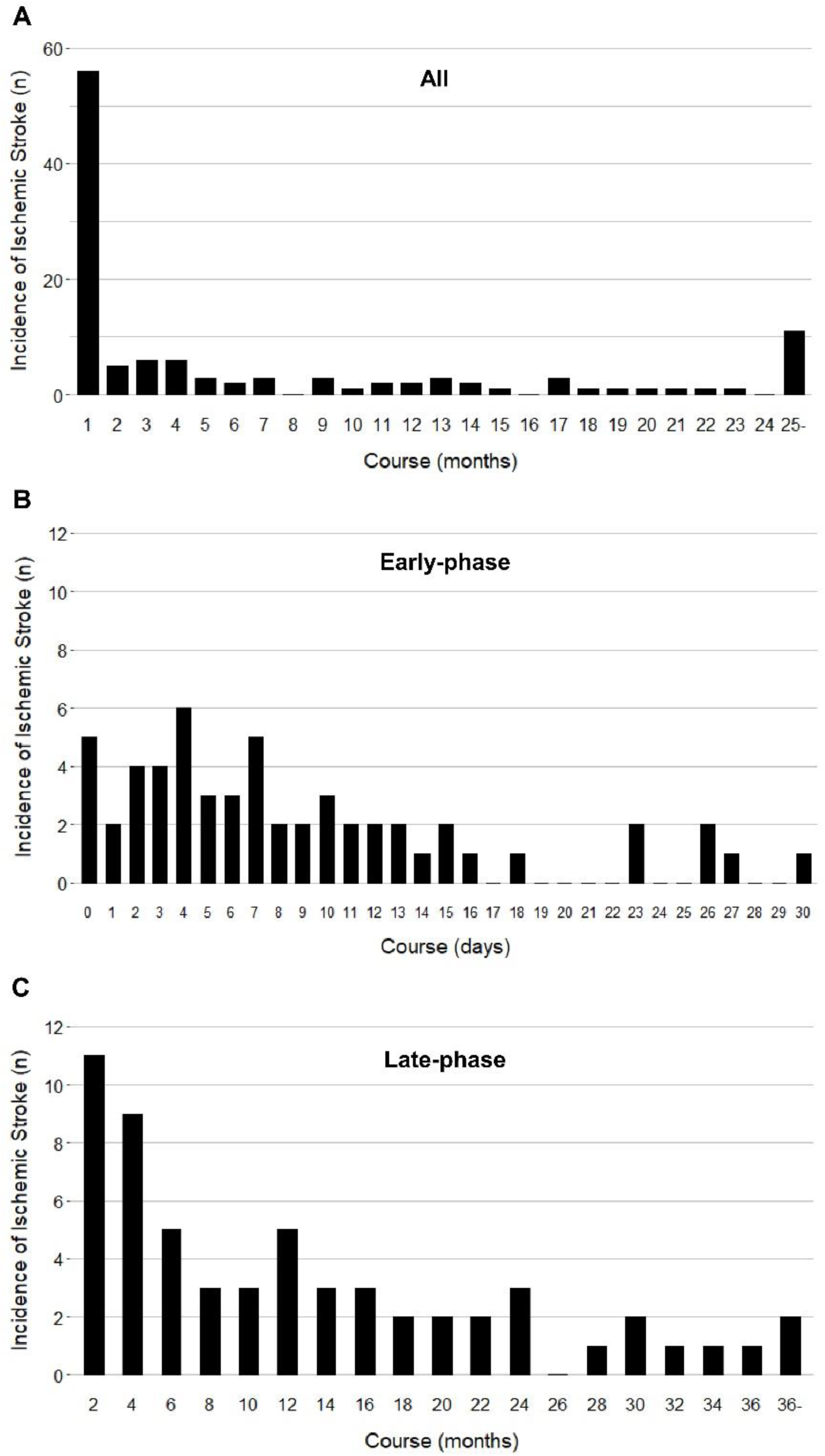
Temporal trends in the incidence of ischemic stroke. All (A), early-phase (B), and late-phase (C) ischemic stroke.

Among underlying causes of IS, cardioembolic stroke was the predominant subtype in both early and late phases, followed by large-artery atherosclerotic stroke (**Figure 2A**). In patients with AF, the occurrence of cardioembolic stroke was consistent across different time points. However, in patients with sinus rhythm, cardioembolic stroke more commonly occurred during the early phase, while the proportion of atherosclerotic stroke increased in the late phase (**Figure 2B and 2C, Figure S2**).

**Figure 2.**
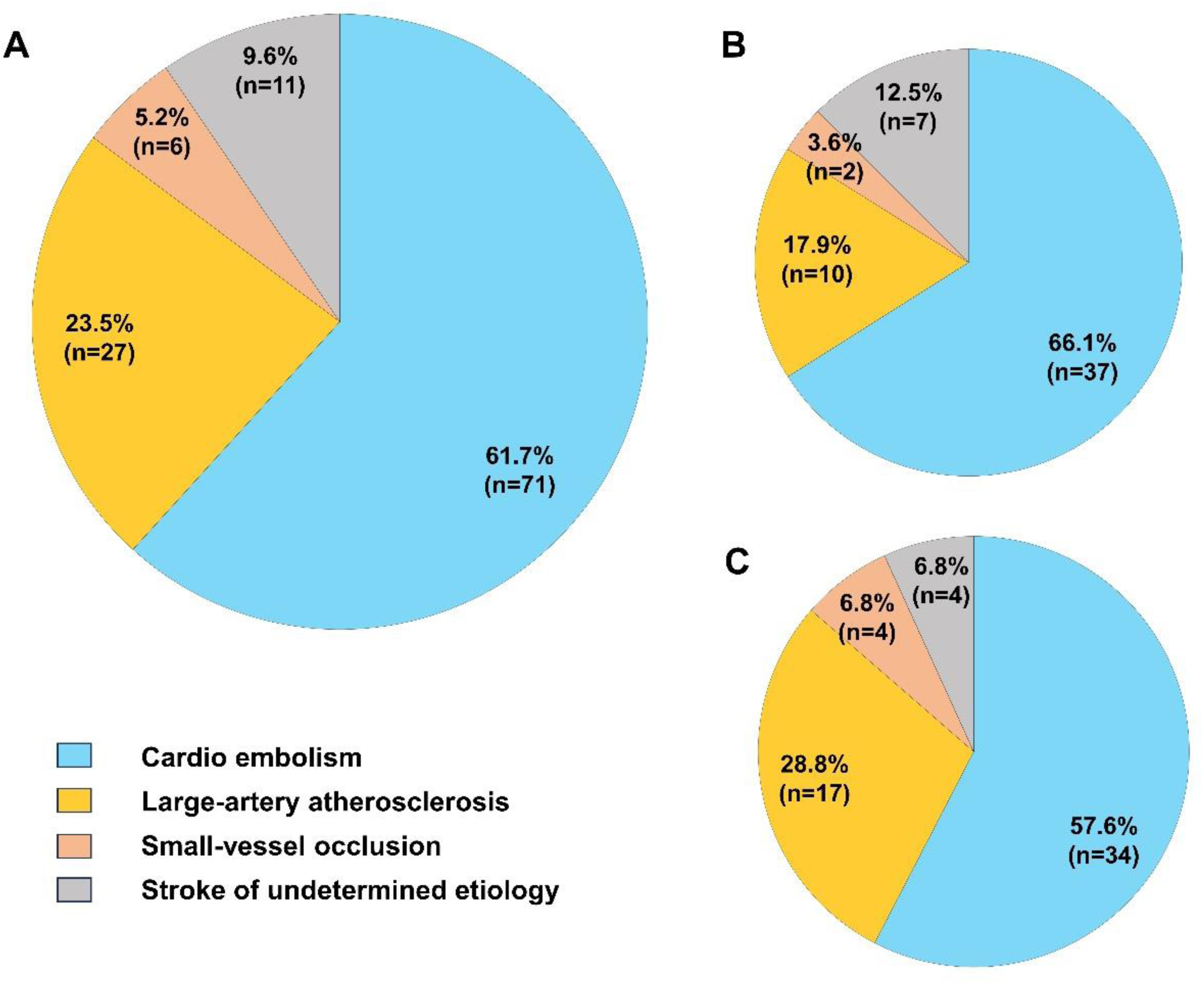
Ischemic stroke subtype. All (A), early-phase (B), and late-phase (C) ischemic stroke.

### Associations of hemoglobin levels and ischemic stroke

For overall IS analyses, the results from Fine–Gray models accounting for the competing risk of death showed that hemoglobin levels (i.e., anemia or not), as well as other risk factors, were not significantly associated with IS incidence (adjusted sHR 1.14; 95% CI 0.75–1.72, p=0.54), except that prior stroke was associated with IS incidence (sHR 1.69, 95%CI 1.05–2.74, p=0.03) (**Table S2**). Notably, the stratified analysis by onset phases revealed distinct associations. High hemoglobin levels (non-anemia) were independently associated with an increased risk of early-phase IS (adjusted sHR 2.05; 95% CI 1.09–3.83, p=0.03). In contrast, the Cox model with time-varying covariates demonstrated that hemoglobin levels were not associated with late-phase IS risk (adjusted HR 1.21; 95% CI 0.65–2.23, p=0.55) (**Table 2**).

**Table 2.**
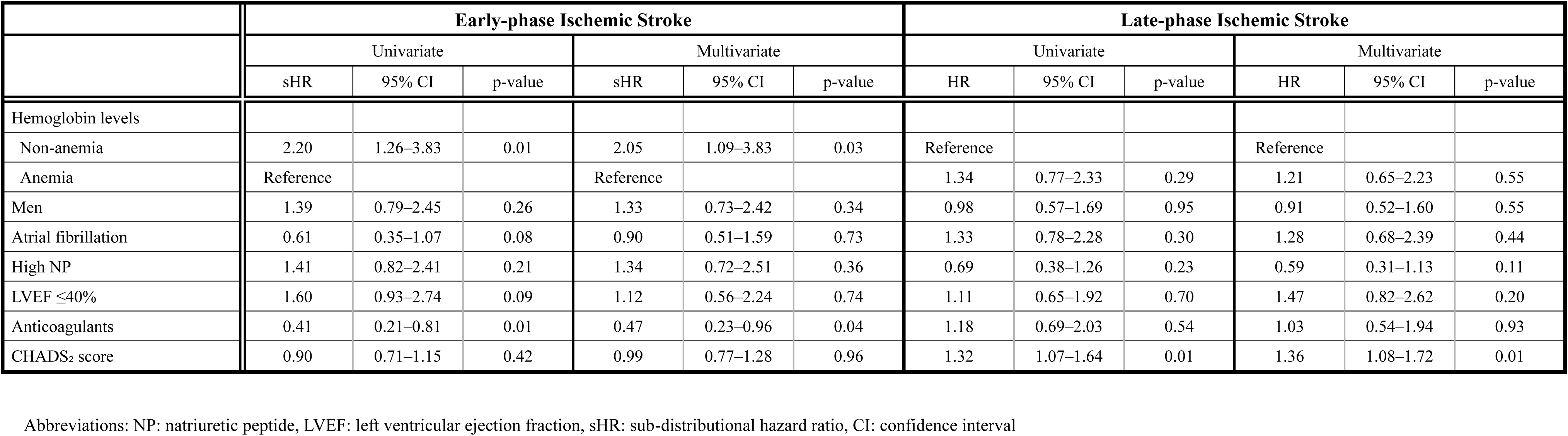
Adjusted and unadjusted Fine and Gray models (early-phase) and Time-varying Cox proportional hazard models (late-phase) for ischemic stroke

To further evaluate and illustrate the non-linear relationship between hemoglobin levels and IS risk, restricted cubic spline analyses were performed. **Figure 3** showed the temporal variations in baseline hemoglobin levels and their association with IS risk. In the early-phase IS group, the relationship between hemoglobin and IS risk was significantly observed at levels above 12.0 g/dL. Conversely, in the late-phase IS group, the relationship between hemoglobin levels and late-phase IS risk showed different findings compared to the early-phase, with no significant association.

**Figure 3.**
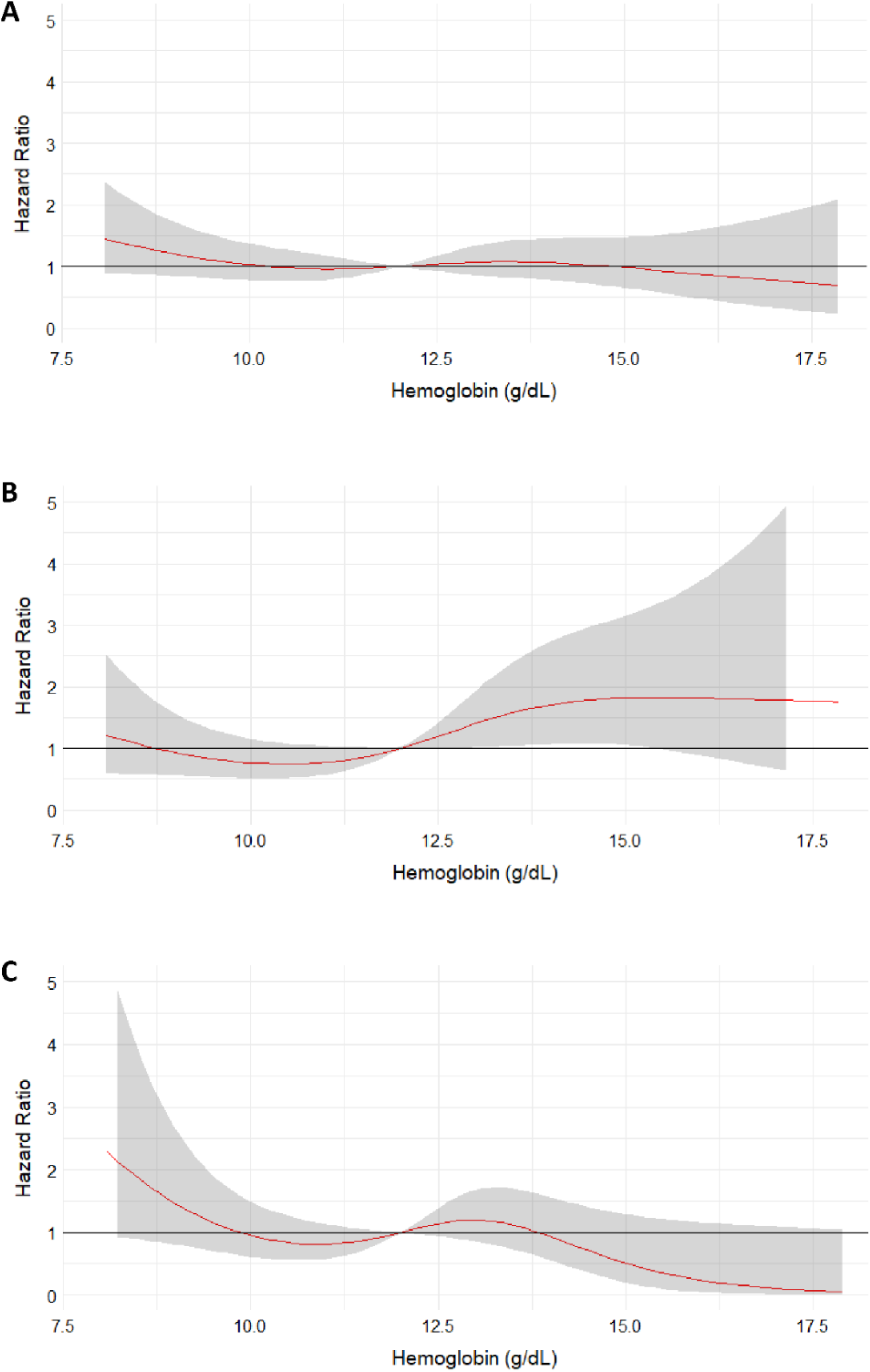
Association between hemoglobin and ischemic stroke risk in patients with ADHF. Restricted cubic spline curve for ischemic stroke risk according to all (A), the early-phase (B), and the late-phase ischemic stroke (C).

### Subgroup analyses for LVEF and AF

In the LVEF-stratified analysis (>40% vs, ≤40%), adjusted Fine–Gray models indicated that high hemoglobin levels were significantly associated with an increased risk of early-phase IS in patients with LVEF >40% (adjusted sHR 3.43; 95% CI 1.44–8.19, p=0.006) (**Table S3**), while in patients with LVEF ≤40%, high hemoglobin levels were not independently associated with early-phase IS (adjusted sHR 1.15, 95% CI 0.55–2.40, p=0.71) (**Table S4**). For late-phase IS, hemoglobin levels were not consistently associated with IS risk in patients with LVEF >40% (HR 1.15, 95%CI 0.50–2.63, p=0.74) (**Table S3**) and those with LVEF ≤40% (HR 1.11, 95%CI 0.47–2.63, p=0.81) **(Table S4**). These associations were visually supported by restricted cubic spline curves (**Figure S3**).

In the subgroup analysis stratified by the presence or absence of AF, adjusted Fine– Gray models for non-AF patients indicated that high hemoglobin levels were significantly associated with early-phase IS (adjusted sHR 2.24, 95% CI 1.10–4.55, p=0.03) (**Table S5**). Similarly, a trend toward increased early-phase IS risk was observed in patients with AF, although this finding did not reach statistical significance (adjusted sHR 1.93, 95% CI 0.78– 4.77, p=0.15) (**Table S6**). In late-phase IS, hemoglobin levels were not associated with IS risk in either group (non-AF: HR 0.69, 95% CI 0.29–1.64, p=0.41; AF: HR 1.82, 95% CI 0.81–4.09, p=0.15) (**Table S5** and **S6**). These results were illustrated using restricted cubic spline curves (**Figure S4**)

## Discussion

Using a nationally representative cohort of ADHF patients in Japan, this study found several important information: (1) IS following ADHF hospitalization frequently occurred at an annual rate of 3.6% per person, with approximately half of events occurring within the first 30 days, and the median time to onset was 7 days in early phase groups; (2) cardioembolic stroke was the predominant cause of IS across both early and late phases; (3) there are significant differences in the patient characteristics between the early and late IS groups, with particular risk factors prevalent in each phase; and (4) high hemoglobin levels at hospitalization were significantly associated with an increased risk of early-phase IS, a relationship that consistently maintained in sensitivity analyses.

This study identified higher hemoglobin levels as a significant risk factor for early-phase IS after worsening HF episodes, a finding that aligns with previous clinical observations.^31–33^ Elevated hemoglobin raises blood viscosity, promoting thrombosis,^34^ and can impair endothelial function, increasing peripheral vascular resistance and potentially lowering cerebral perfusion.^35^ Additionally, high hemoglobin levels lead to platelet activation through the release of adenosine diphosphate, further promoting a thrombotic environment.^36^ These findings suggest that higher hemoglobin levels in ADHF patients warrant careful monitoring for the prevention of early-phase IS. Regardless of the presence of AF, cardioembolic strokes are more prevalent in the early-phase IS, supporting the fact that HF itself fulfills the components of the Virchow triad: blood stasis, endothelial dysfunction, and hypercoagulability, creating a condition conducive to the formation of intracardiac thrombi.^37^ Elevated hemoglobin levels, combined with significant hemodynamic fluctuations during the early phase of ADHF, are likely to contribute to enhanced blood coagulability. To prevent IS events in the vulnerable periods after ADHF hospitalization, antithrombotic therapies may be useful as anticoagulant usage positively associated with reduced risk of IS in the multivariable models.

In the late-phase analysis, a higher CHADS₂ score was associated with late IS, as an established factor. Notably, there was no association between hemoglobin levels and late-phase IS; however, restricted cubic spline curve demonstrated that lower hemoglobin levels would be risk of late-phase IS in AF and regardless of LVEF. Furthermore, in the patients with LVEF ≤40%, the restricted cubic spline showed U-shaped curve, suggesting lower hemoglobin levels also may contribute to an increased risk of early-phase IS. Associations of anemia with increased IS risk has been documented in non-HF populations,^31,32^ likely due to reduced oxygen delivery to tissues and compromised cerebral vascular regulation.^38,39^ Additionally, hyperdynamic circulation in anemic states can promote IS risk through increased endothelial adhesion molecule expression and inflammation.^40^ In cases of iron deficiency anemia, elevated erythropoietin levels can further elevate thrombosis risk through erythrocytosis and thrombocytosis.^41^ Collectively, relationships between hemoglobin levels and IS appear nuanced in patients with HF, influenced by phase-specific variations in risk. Notably, the restricted cubic spline analysis revealed distinct temporal patterns between hemoglobin levels and IS risk across the early and late phases. These findings underscore the complexity of the impact of hemoglobin levels on IS risk in HF populations, emphasizing the need for time-sensitive management approaches. This study identifies a critical high-risk period for IS following ADHF hospitalization and highlights cardioembolic stroke as a substantial concern, even in ADHF patients without AF. These findings suggest a potential role for prophylactic antithrombotic therapy in high-risk subgroups. However, despite promising theoretical benefits, recent randomized controlled trials have not yet confirmed a net benefit of prophylactic anticoagulation in HF patients without AF,^3,42,43^ largely due to the heightened risk of bleeding complications, leaving the benefit-risk balance uncertain. To refine this strategy, recent proposals suggest implementing targeted anticoagulation strategies for high-risk patients during periods of peak risk.^13,15,29,30,44,45^ It may also be critical to focus on minimizing exposure to anticoagulants in lower-risk intervals.^4^ Our findings align with this approach, identifying the initial two weeks post-ADHF hospitalization as the highest risk window, with a median time to IS onset of 7–10 days post-ADHF hospitalization.^2^ Anemia is a well-known risk factor for bleeding complications in patients receiving antiplatelet or anticoagulant therapy;^46–48^ conversely, patients with high hemoglobin levels—a group at lower bleeding risk—may particularly benefit from more targeted anticoagulation. By identifying specific risk factors and temporal patterns of IS, this study adds valuable insights to inform future randomized controlled trials aimed at refining anticoagulation strategies during the early phase after ADHF.

This study had several strengths. First, we elucidated the subtypes of IS associated with ADHF, which is critical for the choice of appropriate antithrombotic therapy. In addition, our registry designed in an all-comer style minimizes selection bias. However, there were several limitations in the present study. First, it is subject to unmeasured confounders, limiting our ability to establish causation. Additionally, we did not specify anemia subtypes (i.e., iron deficiency anemia, etc), which may have different effects on IS risk. We also lacked data regarding intravenous administration of heparin during hospitalization. Finally, the association of anemia with IS is well-documented in non-HF populations, while Cox proportional hazards models incorporating time-varying hemoglobin levels showed no significant association with late-phase IS risk. We did not track detailed hemoglobin trends beyond three time points, limiting our ability to comprehensively assess the relationship between hemoglobin levels and late-phase IS risk.

## Conclusions

In a cohort of ADHF patients with long-term follow-up, we identified a higher incidence of IS within the first 30 days after hospitalization. The relationship between hemoglobin levels and IS risk varies by phase, suggesting that non-anemia might serve as a potential marker for high-risk patients in the early phase. Incorporating hemoglobin levels into risk stratification could help targeted screening and preventive strategies to reduce these events.

## Data Availability

We confirm that all data referred to in the manuscript are available upon request.

## Acknowledgment

None.

## Central illustration

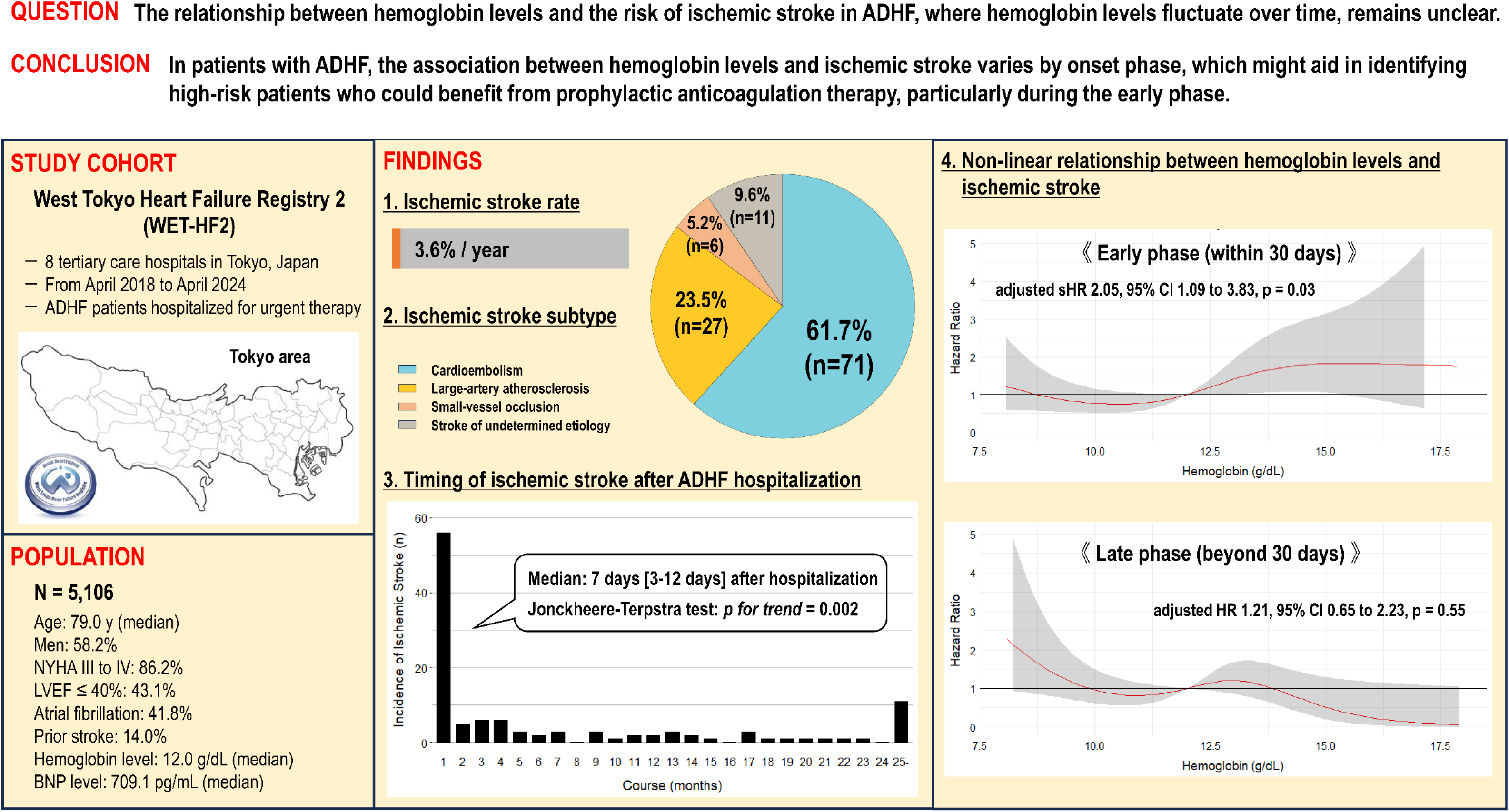

